# A health systems analysis of non-compliance to the maternity referral system by health facilities in Uganda: Findings from a mixed-methods study

**DOI:** 10.1101/2024.04.08.24305473

**Authors:** Henry Zakumumpa, Caroline Kyozira, Martin Bulamu, Remmy Buhuguru

## Abstract

**Introduction:** A fully functional referral system is central to achieving quality maternal and newborn care. In 2018, Uganda established Kawempe National Referral hospital (KRNH) as the highest level of care for maternity services. There is a dearth of data examining if the referral function for which it was established has been met. The objectives of this study were two fold; i) to assess the appropriateness of in-coming referrals at KRNH ii) to examine factors underpinning referral decisions at lower referring facilities across Uganda.

**Methods:** This was a two-phased, mixed-methods study involving 76 health facilities across Uganda. In Phase One, we conducted a cross-sectional study examining incoming referrals at KNRH between July 2019 and August 2020 based on routinely collected data in the Health Information System (HMIS) in-patient registers. In Phase Two, we qualitatively explored factors underpinning maternity referrals at 75 purposively selected facilities. Descriptive statistics were managed in STATA (v14) while qualitative data were analyzed by thematic approach.

**Results:** A total of 461,226 referrals were made to KNRH during the period under review. The three most common indications for referral at KNRH were; previous caesarian scar, premature rupture of membranes and obstructed labour. Facilities mandated to offer comprehensive emergency obstetric care such as general hospitals (84%) and sub-district hospitals or Health Centre IVs (18%) were the two leading sources of referrals at KNRH. Shortage of basic commodities, poorly equipped theatres, deficiencies in skills for labour, study leaves for maternity workforce at lower facilities contributed to unnecessary referrals.

**Conclusion:** Several of the indications of referrals at KNRH are ideally manageable at lower referring facilities such as district and sub-district hospitals levels suggesting deficiencies in the national obstetric referral system. Interventions for strengthening basic clinical skills in the management of labour at lower referring facilities and addressing underpinning health system constraints is recommended.

## BACKGROUND

Reducing maternal mortality is a global health priority [1], [2]. sub-Saharan Africa (SSA) has the highest maternal mortality rate in the world [3]. Estimates of the maternal mortality ratio (MMR) for SSA, as a region, have been as high as 600 per 100,000 live births [4]. The Sustainable Development Goals (SDGs) have set targets for reducing maternal mortality in SSA and the rest of the globe [1], [2].

Health system constraints such as low skilled birth attendance and the stock outs of commodities have been identified as influential on maternal health outcomes in SSA [5]. However, cultural and socio-economic factors also play a role [6]. More direct causes of maternal mortality include hemorrhage, unsafe abortion, hypertensive disorders in pregnancy, obstructed labour and sepsis [7]. Emergency obstetric care(EmOC) is a priority intervention for reducing maternal mortality [8]. Emergency obstetric care refers to care given in the event of complications during and after labour that pose a threat to the life of the mother or child [8]. Within the suite of interventions under EmOC, referrals for more advanced care for mothers in labour is a central tenet underpinning the quality of maternity care provided [9]. Referrals are important in that they allow for advanced specialist handling of complications of females in labour before they pose a danger to the life of the mother and the unborn child [10]. The WHO recognizes functional referral systems as a key indicator of the quality of maternal care [11] (**Figure 1**).

In SSA, several countries such as Ethiopia have set up specialized referral hospitals for EmOC [12]. Although, there is an accumulating evidence base on understanding maternal care services at the primary-care level of service delivery in SSA [13]-[15], there is a paucity of evidence on the uptake of maternity services at newly established specialized national referral hospitals [16]. These data are important in understanding trends in health seeking for obstetric health care and the underpinning health system constraints. This is critical for health services planning and in devising appropriate interventions for achieving targets for reducing maternal mortality in SSA [1], [2].

Uganda has a maternal mortality ratio of 343 out of 100,000 live births [17], [18]. An estimated 2% of women in Uganda die due to maternal causes [17], [18].

In 2018, Uganda established the Kawempe National Referral Hospital (KNRH) as a specialized national centre of excellence in maternal and newborn care services. The hospital was intended to handle referrals from lower-level facilities that require more advanced and specialized care [19]. The Ugandan health system has a hierarchical referral system (**Figure 2**) where primary care level facilities (such as sub-county health centre IIIs) are expected to provide basic EmOC while sub-district and district hospitals are expected to offer comprehensive EmOC while more advanced cases are referred to the tertiary level of care and ultimately to the national referral hospital along an apex cascade chain [8].

Although Kawempe National Referral hospital has been in operation since 2018, there is a dearth of research reporting trends in the uptake of lifesaving services at KNRH, the portfolio of patients it serves since its inception and, if the referral function for which it was established is being met. The objectives of this study were two fold; i) to assess the appropriateness of in-coming referrals at KRNH ii) to examine factors underpinning referral decisions at lower-level referring facilities across Uganda.

## MATERIAL AND METHODS

### Study design

This was a mixed-methods sequential explanatory research design [18]. The study was conducted in two phases, implemented sequentially [18]. In Phase One were conducted a cross sectional study of incoming obstetric referrals at Kawempe National Referral Hospital in Uganda between 1^st^ July 2019 and 31^st^ August 2020. In Phase two, we qualitatively explored factors underpinning referral decision-making at 75 lower referring facilities across Uganda.

### Study sites

#### Phase One

Kawempe National Referral Hospital (KNRH) is located in one the administrative units of the Ugandan capital city of Kampala in central Uganda. The hospital was established in 2018 as a specialized national referral hospital for maternity care. KNRH is the highest level in the maternity referral chain in Uganda. With over 32,000 births per year, the hospital is also the largest and busiest specialized maternity care facility in Uganda with 80-100 deliveries per day [20].

#### Phase Two

We purposively sampled at total of 15 facilities with the highest number of referrals at KNRH to qualitatively explore factors underpinning the high number of referrals. We selected five facilities from each of three levels of care in the Ugandan health system (**Figure 2**); at the level of i) General hospitals, ii) sub-district facilities (Health Centre IV) and iii) sub-county health facilities (Health Centre IIIs). For regional representation, we selected five facilities from each of four geographic regions of Uganda (Northern, Western, Eastern and Southern). In each of the four regions, we selected; a) one general hospital (including two private ones) b) two Health centre IVs c) three Health centre IIIs. We aimed for an in-depth exploration of factors influencing referral decision-making at lower referring facilities outside of central Uganda (which contributes the highest number of referrals).

### Data collection

#### Phase One

Data were collected between 1st July 2019 and 31st August 2020 at Kawempe National Referral Hospital (KNRH).

Data were also extracted from three principal sources; a) from routinely collected maternity services records from the Health Information System (HMIS) In-patient Registers, b) anonymized patient-level delivery data were extracted from maternity registers maintained at the hospital and c) from the District Health Information System (DHIS2). Due to a decentralized filing system where each department keeps its own, largely manual system, data were extracted from a number of primary records from individual units at KNRH. We included all females referred with pregnancy-related complications while pregnant (antenatal complications needing urgent delivery), during labour or within immediate post-partum.

Table 1 shows the list of records reviewed for this study.

**Table 1:**
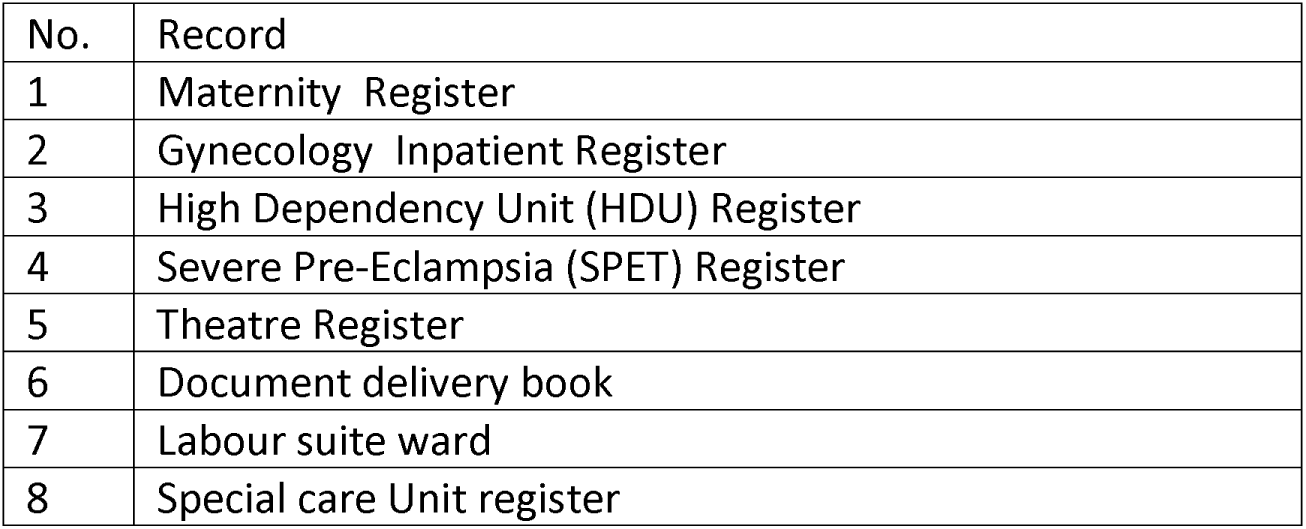
Primary records from which data were extracted.

From the described registers, we extracted; i) Unique patient identifiers, ii) Date of admission, iii) Age of client, iv) Reason for referral and v) Referring facility.

#### Phase Two

We conducted semi-structured interviews (SSIs) with facility in-charges of the selected health facilities to explore the health system context(s) underpinning referral decision-making themed around the WHOs’ six building blocks of the health system (supplies and medicines, human resources for health, financing, governance, service delivery and health information systems). A pre-tested interview guide was used constructed around the six ‘building blocks’ of a health system. Interviews were conducted by the first author with the support of four research assistants experienced in health services research. On average the interviews lasted between 45 to 60 minutes. Face-to-face interviews were conducted in participants’ offices at the 75 purposively selected facilities between June and September 2021.

### Ethical clearance

This research received ethical approval from Mildmay Uganda Research Ethics Committee (MUREC) under instrument: REC REF 0712–2020. MUREC is accredited by the Uganda National Council of Science and Technology (UNCST). All participants provided oral consent form before participating in the study which was witnessed by their facility in-charges. All patient data from Kawempe National Referral Hospital were fully anonymized before being accessed by the investigators.

### Data analysis

#### Phase One

Extracted data were cleaned, edited and initially entered into EpiData version (version 3.1) (EpiData Association, http://www.epidata.dk/). Data were subsequently exported into STATA (version 14) (College Station, TX, USA) for descriptive analyses that included frequency counts and percent distributions.

#### Phase Two

We followed the processes recommended for ensuring rigour qualitative data analysis suggested by Miles & Huberman (1990). We made audio recordings of all of the interviews and then transcribed each interview verbatim. In terms of data analysis procedures, we followed four major steps. However, this was a largely iterative process. The first step involved *data familiarization* through multiple readings of interview transcripts. The second step entailed *generating a coding framework.* Codes were inductively generated from the interview transcripts. The developed coding scheme was applied to the interview transcripts. The third stage was that of *abstracting the coded data into thematic categories* based on the six ‘building blocks’ of the health system (supplies and medicines, human resources for health, financing, governance, service delivery and health information systems). The emergent inductive or data-driven codes were then grouped under six thematic matrices. The fourth and final step was that of *Interpretation and overall synthesis* [36] which involved all the authors.

## RESULTS

### Descriptive characteristics

During the twelve months of the study, there were a total of 461225 referrals to Kawempe National Referral Hospital.

#### Mode of delivery

In terms of mode of delivery (cesarean section / spontaneous vaginal delivery), **Table 2** shows that the mode of delivery was even across most of the age categories at KNRH. However, for the relatively younger age groups of 15-19 and 20-24, delivery via spontaneous vaginal delivery was significantly higher than through cesarean section. **Figure 3** shows that almost a half of all cesarean deliveries at Kawempe National Referral Hospital were for mothers between 25 and 35 years of age.

**Table 2:** Trends in mode of delivery at KNRH

| Age categories | Mode of Delivery |  |
| --- | --- | --- |
|  | C/S | SVD |
| Children (<14 Years) | 8 | 8 |
| Adults (36 - 60 Years) | 518 | 511 |
| Adolescents (15 - 19 Years) | 888 | 1214 |
| Young Mothers (20 - 24 Years) | 2948 | 3463 |
| Youths (25 -35 Years) | 4127 | 4174 |
| *Key: C/S: cesarean section, SVD: spontaneous vaginal delivery* |  |  |

#### Referrals at KNRH by age group

**Table 3** shows that more than a half of all referrals to Kawempe National Referral Hospital (KNRH) were of women between the 25-35 age range. Young mothers (20-24 years) were the second most represented age group in incoming referrals at KNRH. Adolescents account for more than 8% of all referrals to KNRN while a total of 224 children (<14 Years) were referred to KNRH as obstetric emergencies during the period under review.

**Table 3:** Age range of referrals to KNRH

| Age categories | *n* | % |
| --- | --- | --- |
| Children (<14 yrs) | 224 | 0.05 |
| Adolescents (15 - 19 yrs) | 38444 | 8.33 |
| Young Mothers (20 - 24 yrs) | 142230 | 30.83 |
| Youths (25 -35 yrs) | 240750 | 52.98 |
| Adults (36 - 60 yrs) | 39576 | 8.58 |
|  | 461224 | 100 |

#### Indications of referrals at KNRH

Figure 4 shows the ten most common indications for referrals at KNRH between July 2019 and August 2020. The three most cited indications of referral at KNRH are; previous caesarian section scar, followed by obstructed labour and premature rupture of membranes at term.

Figure 4 also suggests that a number of the top 10 reasons of referrals to KNRH are for conditions which ordinarily can be managed at Health centre IVs and general hospitals (e.g. hypertension in pregnancy, pre/eclampsia) which are mandated to offer comprehensive emergency obstetric care and are ideally staffed with the requisite cadres of health workers for the purpose.

### Indications of referral by age group

**Table 3** shows that for adolescents (15-19 year olds) obstructed labor was the most common reason for referrals to KNRH.

Overall, previous caesarian section scar was the leading indication of referral to KNRH across three age categories; a) young mothers (20-24 year olds), b) youth (25-35 year olds) and c) adults (36-60 year olds).

Obstructed labour was another prominent reason for referrals at KNRH and was consistently so across all age categories (except for those 14 years or younger).

**Table 4** shows that the three most referred indications at KNRH are; previous cesarean section scar, obstructed labor and rupture of membrane at term. These indications of referral were common across all age groups. The notable exception was with regard to children (<14 Years) seeking care at KNRH for whom; pre-term labor, pre-eclampsia and antepartum hemorrhage (APH) were the most frequent indications of referral at KNRH.

**TABLE 4:** MOST REFERRED CONDITIONS BY AGE GROUP

| Reason for referral | No. Referred | Children (<14 yrs) (%) | Adolescents (15 - 19 yrs) (%) | Young Mothers (20 - 24 yrs) (%) | Youths (25 -35 yrs) (%) | Adults (36 - 60 yrs) (%) |
| --- | --- | --- | --- | --- | --- | --- |
| In Labor | 209149 | 168(0.1) | 17817(8.5) | 64182(30.7) | 108573(51.9) | 18409(8.8) |
| Previous Scar | 53779 | 0(0) | 916(1.7) | 12273(22.8) | 35229(65.5) | 5361(10) |
| Obstructed Labor | 26525 | 0(0) | 3469(13.1) | 9615(36.3) | 11857(44.7) | 1584(6) |
| Premature rupture of membranes | 15783 | 0(0) | 1238(7.8) | 5676(36) | 7657(48.5) | 1212(7.7) |
| Preterm Labor | 14437 | 14(0.1) | 1928(13.4) | 5081(35.2) | 6384(44.2) | 1030(7.1) |
| Malpresentation/Malposition | 14172 | 0(0) | 1134(8) | 4412(31.1) | 7029(49.6) | 1597(11.3) |
| Hypertension in Pregnancy | 12455 | 0(0) | 667(5.4) | 3485(28) | 6620(53.2) | 1683(13.5) |
| CPD (Cephalopelvic Disproportion) | 11855 | 0(0) | 2438(20.6) | 5221(44) | 3812(32.2) | 384(3.2) |
| Pre-eclampsia/Eclampsia | 11632 | 14(0.1) | 917(7.9) | 3427(29.5) | 5863(50.4) | 1411(12.1) |
| APH (antepartum hemorrhage) | 10165 | 14(0.1) | 604(5.9) | 3025(29.8) | 5567(54.8) | 955(9.4) |
| Fetal Distress | 9006 | 0(0) | 975(10.8) | 3644(40.5) | 4120(45.8) | 267(3) |

#### Characteristics of referring facilities

Uganda runs a hierarchical referral system with lower-level primary care facilities at the bottom of the pyramid which then refer cases to higher-tier facilities at the secondary level of service delivery through to tertiary hospitals up the ladder with national referral hospitals at the apex as illustrated in Figure 1.

Figure 5 shows that general hospitals (84%) were the leading sources of incoming referrals at KNRH. This was followed by health centre IVs (18%). Both general hospitals and Health centre IVs are mandated to offer comprehensive emergency obstetric care in the Ugandan health system.

In **Table 4** we show the five leading referring facility categories and the most referred indication within each of those facilities within category of level of service delivery in the Ugandan health system.

**Table 4:**
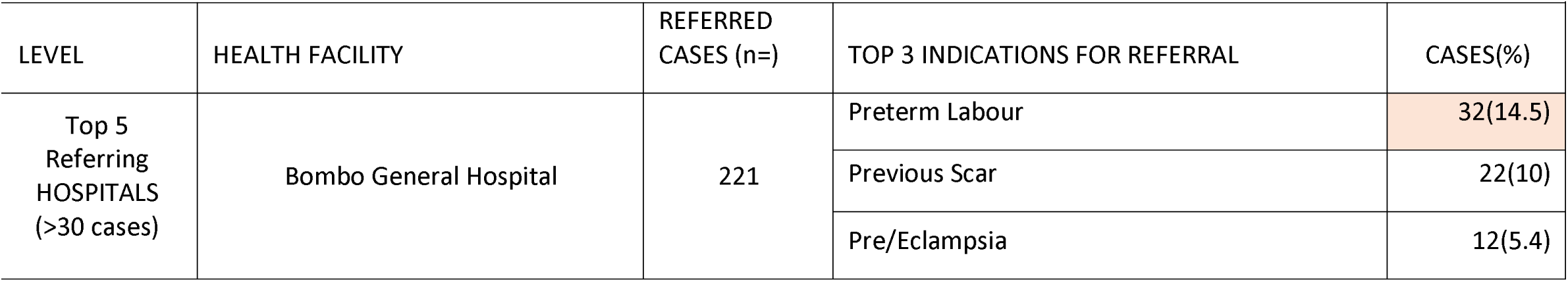
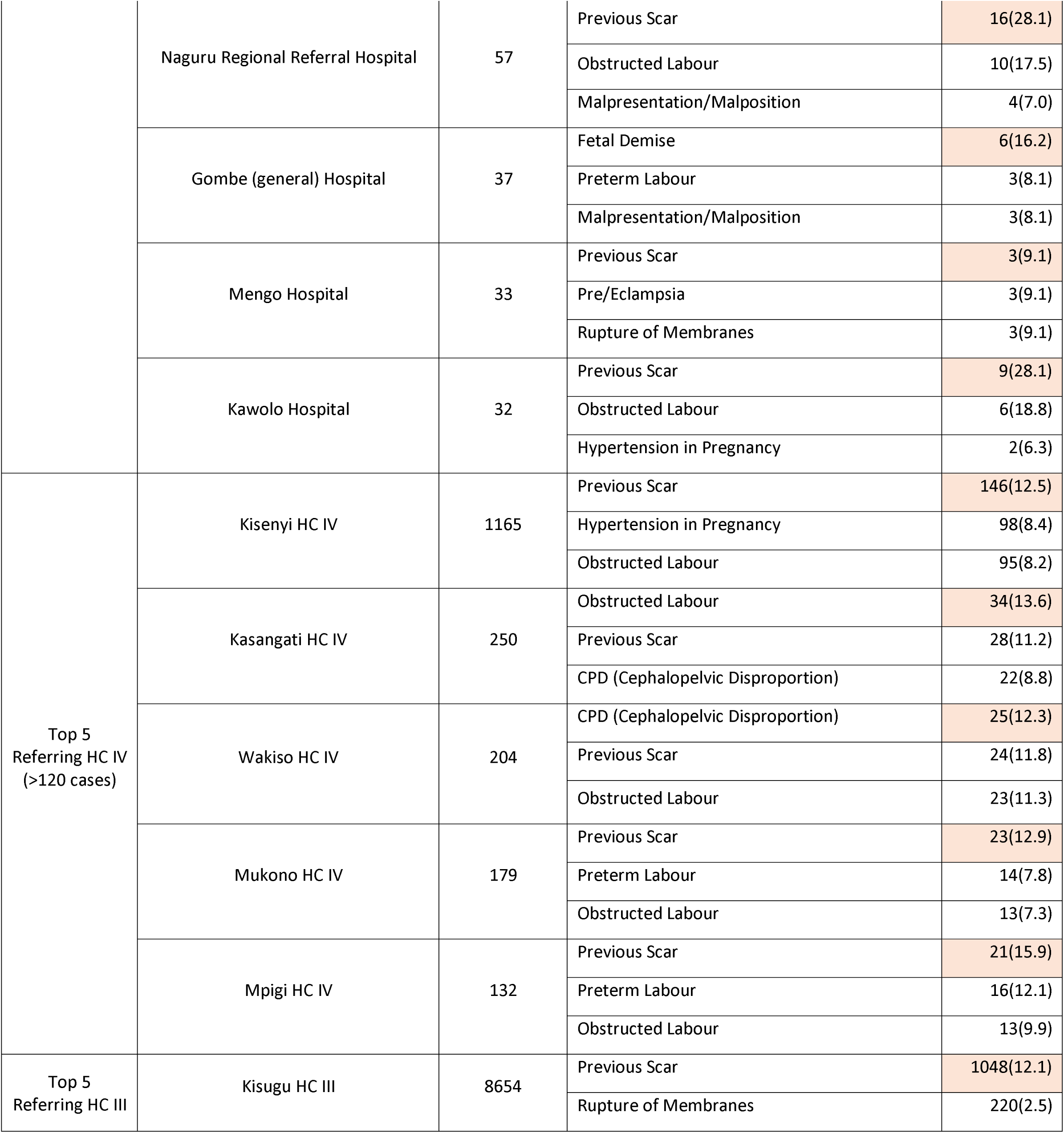
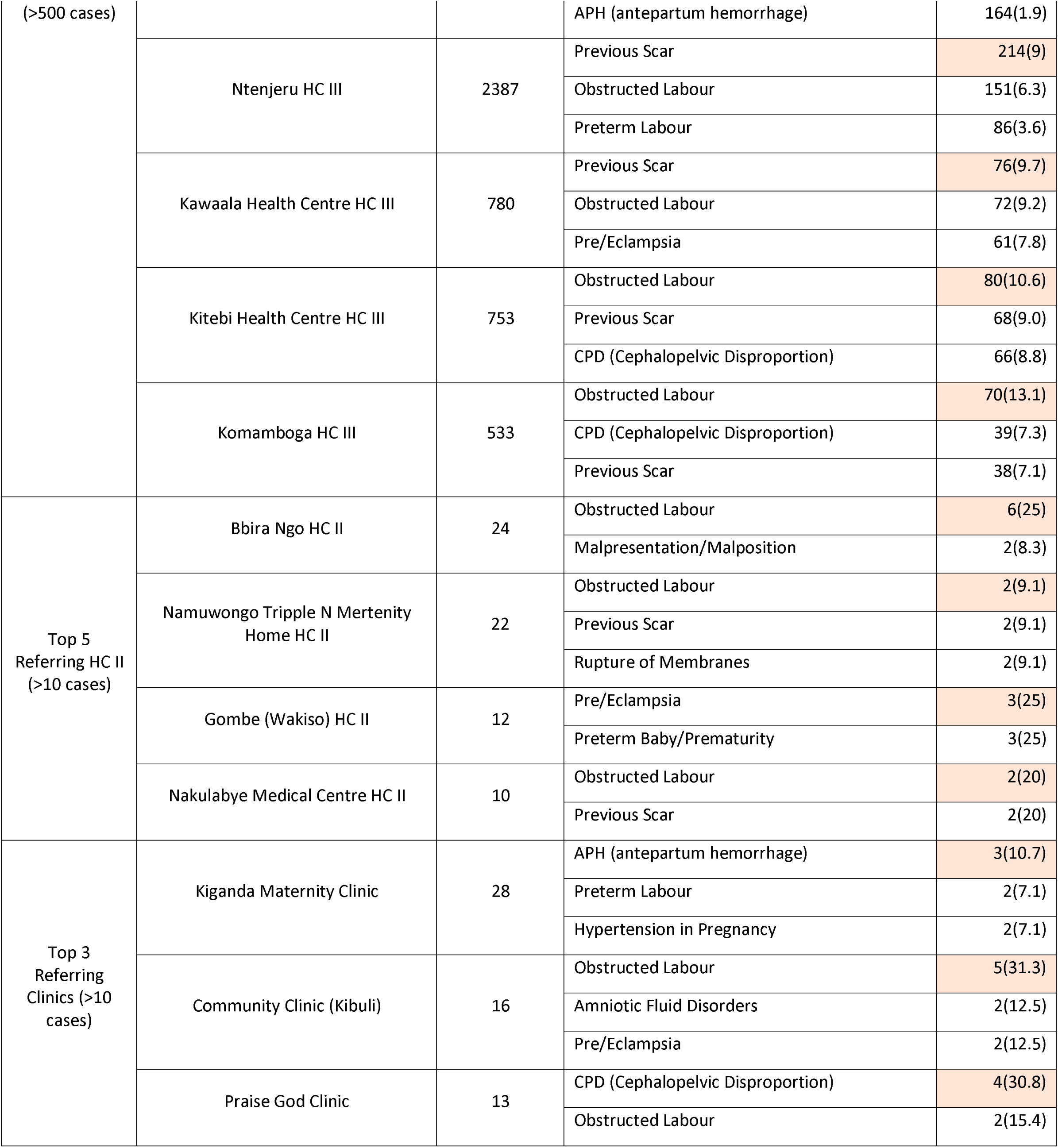
Facilities with the most referrals by level of service delivery

Two of the top five referring facilities are at the level of Regional Referral Hospitals (Naguru Hospital and Mengo Hospital). Several of the indications of referral to KNRH are ordinarily manageable at the Regional Referral hospitals which have obstetrics and gynecology specialists and the requisite infrastructure to handle the cases referred to KNRH (such as previous scar).

From Table 4 it can be discerned that Previous caesarian section scar was the most common referred condition across the top referring facilities within the two category of facilities (hospitals and health centre IVs).

Obstructed labour was the second most referred to obstetric emergency in health centre IVs. For the small clinics, the obstetric emergencies referred to KNRH were more diverse compared to higher-tier hospitals.

### Geographical distribution of in-coming referrals at KNRH

**Table 4** shows that the majority of referrals to KNRH were from the capital city of Kampala, where the hospital is based, and the neighboring districts of Wakiso and Mukono.

The majority of referrals were from public municipal satellite clinics of Kampala City Authority (KCCA) (41%) and from the neighboring district of Wakiso (21%). Overall, 61% of all incoming referrals at Kawempe National Referral, during the period under review, are from only 2 (out of 135) districts of Uganda.

## SECTION B: Factors underpinning referrals at lower referring facilities

### Human resources for Health

#### Deficiencies in skill sets for labour

Interviews with health workers revealed deficiencies in skills sets for labour at the fifteen facilities with the highest number of referrals at KNRH (**Table 4**). Specific skills gaps were identified by health workers who highlighted the need for training on monitoring of labour such as the use of the partograph (or labour chart) that are required as part of routine maternity services provision and are helpful in screening for complications in pregnancy. Participants were emphatic in relaying the notion that routine skills training and refresher courses are sorely needed at lower referring facilities especially around emergency obstetric care. Management of pregnancy-induced hypertension was frequently mentioned as an area that needed continuous medical education as many of the referrals at KRNH were with regard to hypertension in pregnancy that stemmed from poor management in the earlier trimesters of pregnancy such as high-risk expectant mothers not being given anti-hypertension medication. The monitoring of vital signs in expectant mothers was also flagged as a weak area in the fifteen facilities with the highest referrals.

#### Shortages of physicians

Rural-based sub-district facilities reported shortages of physicians to conduct surgical procedures even in cases where theaters were available. Although sub-district facilities are mandated to have physician cadre on their staff (Figure 2), seven of the facilities we visited reported that they did not have a physician on the ground to handle complications in pregnancy or cases such as managing women with previous scars. The shortage of skilled health workers compelled lower referring facilities to recommend referrals even for basic procedures on account of shortage of medical doctors.

*‘You find the whole of the maternity section of Mbarara Regional Referral Hospital is as congested as a village market. Do you know why? Because doctors at sub-district health centres are not on the ground. The theaters are closed. So you get so many referrals at regional referral hospitals even for things like a cesarean section because doctors are not operating at lower level facilities’ [Facility in-charge, Health Centre IV, Western Uganda, Female]*

A related challenge was the common phenomenon of many of the health workers not being on the ground on account for being on official and non-official study leaves especially those with the requisite skills such as Clinical Officers and physicians especially at rural work stations. Hence, although many of the facilities had health workers with the requisite skills on their staff, many were actually not on the ground.

*‘You find that at a Health Centre IV there are supposed to be two staff, a senior medical officer and a medical officer. Now, if one of them goes on study leave to do a Master of Public Health, the remaining one cannot attend to all the patients in a whole sub-district and the same time do cesarean sections’ [Facility in-charge, Health Centre IV, central Uganda, female]*

### Maternal care commodity shortages

A common refrain across the facilities we visited was the shortage of basic commodities needed in routine maternal care. In Western Uganda, health workers reported that they had not been supplied with commodities from National Medical Stores (NMS), the national medicines supplier for the past three months. Radiology supplies were frequently mentioned as out of stock and yet these are key in diagnosing pregnancies with complications. In such situations, health workers reported that facilities that include general hospitals were compelled to refer expectant mothers to tertiary hospitals which were perceived as better resourced. Basic commodities used in labour such as gloves and syringes were reported to be out of stock in several facilities we visited.

*‘We have not received supplies from National Medical Stores in the last three months. Even when they come, the supplies they give us last a month, instead of two months. So, it means I am always in a constant short fall of drugs I need to serve the number of patients we serve here. The supplies we get is only 50% of what I feel is required’. [Hospital Superintendent, general hospital, Northern Uganda, male]*.

A related constraint reported across multiple facilities was that of shortage of basic equipment needed in maternal care. The frequently mentioned items include basic items such as delivery beds, oxygen concentrators, anesthetic machines, emergency drug trollies and bulb syringes.

The physical state of theaters at primary care facilities was said to be in poor shape and not amenable to conducting obstetric procedures even when the requisite personnel (such as physician cadres) were available. As such, referrals were frequently made higher up the maternity care cascade due to the state of disrepair of theatres at sub-district level in Uganda.

### Financing

#### Insufficient operational grants to facilities

Facility in-charges reported that quarterly funding releases from the central government to public facilities for routine operational expenses dubbed ‘primary health care’ or PHC grants were woefully inadequate to cater for all operational expenses that include repair of basic equipment needed in maternity services such as theaters. A large district hospital in Western Uganda reported that their quarterly release for all hospital expenses (including medical equipment) was as little as 40 million Uganda shillings ($ 6,250). This was in spite of the fact that district hospitals attend to hundreds of patients and burgeoning clients from neighboring districts. In these cash-strapped circumstances, facilities were frequently unable to maintain physical infrastructure and equipment for routine maternal care and opted to refer patients to the tertiary level of care even when this was not clinically warranted.

‘*There is severe under funding. The money we are given as a general hospital is for recurrent expenditure for things like electricity and water, there is nothing for equipment such as X-ray, ultra sound and anesthetic machines. We do not receive any single coin to construct even a toilet! So, how can I buy expensive equipment to refurbish the theatre for cesarean sections? I admit over 500 mothers in maternity every month. There is a cost to every operation of a pregnant mother in terms of gloves, in terms of drugs, disposable gloves, surgical blades, gauze, cotton but I don’t have money for that. All I have to do is refer the cases which need surgeries’ [Hospital superintendent, General Hospital, Western Uganda, male]*

### Gaps in maternity records data sharing

#### Paucity of records on in-coming referrals

Participants across the health facilities we visited frequently mentioned that information sharing between lower referring facilities and facilities receiving in-coming referrals up the higher-tier of the referral ladder was a fundamentally weak (Figure 2). At Kawempe National Referral Hospital it was common to find that patients were received with insufficient background information on indications of referrals and their previous management by lower referring facilities. Uganda runs a health information system (HMIS) incorporating routinely collected data on maternity services however documentation by attending health workers across the cascade was reportedly weak which contributed to low data utilization by referral facilities.

The phenomenon of self-referrals or ‘patient-walk-ins’ was said to be common across the obstetric referral chain in Uganda which further compounded the challenge of paucity of data on in-coming patients at especially the tertiary level of care. Participants acknowledged that the failure by higher-tier hospitals to insist on minimum referral requirements before accepting in-coming referrals such as having basic information on patients’ previous management history from the lower referring facility encouraged the widespread practice of bypassing the established maternity services referral chain in Uganda by both providers and patients.

It emerged that there were no mechanisms for providing feedback to lower referring facilities to enable them learn from the events (such as in strengthening skills in partograph use) which perpetuates non-adherence to the referral chain in Uganda and contributes to accumulating unnecessary burdens at the highest level of maternal care.

### Governance

#### Weaknesses in referral linkages

Our study unearthed gaps in maternity services referral linkages between Regional Referral Hospitals (RRHs) and general Hospitals (Figure 2). Although RRHs are mandated to provide supervisory oversight over general hospitals in the regions under their purview, participants at district hospitals conceded there was a disconnect between the two. It emerged that RRHs frequently don’t perform their supervisory function at general hospitals.

#### Limited support supervisions

On-site supervisions of maternity services at general hospitals by RRHs was said to be dysfunctional. The two levels of care were said to operate independently of each other, in practice, despite the elaborate referral relationships described in written policies and guidelines. This disconnect created an environment where referrals from lower referring facilities to higher-tier hospitals were not rationalized and did not meet the laid down referral criteria and procedures.

*‘Our regional supervisors are supposed to be Regional Referral hospitals but somehow we do not connect in routine operations. The Regional Referral Hospital are supposed to do support supervision in maternity services delivery for lower health facilities but a for a whole year you will never see any person from there come here’ [Hospital superintendent, general hospital, Eastern Uganda, female]*.

## DISCUSSION

A fully functional referral system is central to achieving quality maternal and newborn care in Sub-Saharan Africa [1]. We set out to understand the appropriateness of referrals made to Kawempe National Referral Hospital in Uganda and the factors underpinning referrals decision making at 75 lower referring facilities across Uganda. We reviewed data of maternal and perinatal admissions between July 2019 and August 2020. We found that previous cesarean section scar was the most referred condition at KNRH. Obstructed labour and rupture of membrane at term were the other two most commonly referred conditions at Kawempe National Referral Hospital during the period under review.

Overall, our findings suggest that the ten most common incoming referrals at KNRH stem from initial poor handling of labour conditions at referring lower-level health facilities. These data point to possible deficiencies in basic clinical skills in labour and its management by attending health workers from the primary to the tertiary level of service delivery as well under-resourced health systems and challenging operational contexts at lower-level referring facilities such as the adequacy of the physical infrastructure underpinning maternal care. A study in Ghana by Afari and colleagues [24] found that deficiencies in clinical skills in the management of labour were one of the top four groups of factors impeding maternal care in Ghana. Previous studies have implicated health worker skills in maternal care at primary care facilities [1]-[5].

In this study we found that previous caesarean section scar, premature rupture of membrane and obstructed labor were three leading reasons for referrals at KNRH. A study by Ghardallou and colleagues [20] found that premature rupture of membrane was the second most referred obstetric condition at a tertiary university hospital in Tunisia. Hueston and colleagues [21] found that early obstetric referrals were most likely among women who delivered by cesarean section in a study that pooled data from five states in the United States.

On the whole, our findings suggest that the obstetric care referral system in Uganda is not functioning as intended from the vantage point of the national specialized hospital. More specifically, the majority of the conditions referred to Kawempe National Referral Hospital (such as mothers with previous scars, hypertension in pregnancy) are ordinarily manageable at sub-district level or by district hospitals which have the requisite cadres of health workers (such as physicians and even obstetrics and gynecology specialists, in the latter case) with the competence to effectively manage the referred conditions.

Phase Two of our study implicated health system factors in the non-adherence of referral guidelines at lower referring facilities due to the underpinning operational context such as the shortage of basic commodities for maternal care (such as radiology supplies), severe under funding culminating in sub-optimal physical infrastructure (e.g. poorly equipped theatres) and maternity workforce constraints such as absenteeism. In addition, we found weak referral linkages and communication gaps between lower referring facilities and receiving facilities at the apex of the referral chain in Uganda. Dysfunctional governance regimes where tertiary-level facilities hardly conduct on-site supervision of lower-level facilities in maternity care were identified as contributory factors to the non-adherence to the hierarchical referral system in Uganda (Figure 2). Mbonye and colleagues [8] found that critical staffing gaps at lower-level facilities in Uganda hamper the observance of maternity referral guidelines. In another study from Uganda, Mubiru and colleagues [23] report that women frequently bypass the established obstetric referral chain and that this happens 40-70% of the time. A scoping review found that maternity workforce constraints at the primary care level in developing countries impede adherence to established maternity referral systems [22]. This same scoping review notes that deficiencies in basic communication, the paucity of feedback mechanisms between referrer and receiver were common in maternity referral systems in the developing world.

Together, our findings suggest that KNRH is inundated with ‘normal’ cases that are ideally manageable by less specialized cadres and providers and as such, the national referral hospital is bereft of the occupational experience of managing more specialized cases and its specialists are not being optimally utilized in handling more deserving referrals.

Additionally, the volume of referrals at KNRH which were close to a half a million cases during the period under review, are overwhelming for a single facility. The quality of care at KNPH given the patient load handled and health worker workloads are major points of concern.

In terms of the geographic distribution of incoming referrals, we found that the majority of referrals to Kawempe National Referral Hospital were dominated by peripheral facilities to KNRH from within the Ugandan capital city of Kampala and from three principal neighbouring districts in central Uganda (Wakiso, Mukono and Mpigi). Our data suggests that the profile of incoming referrals at KNRH do not reflect diversity in the geographical sub-regions of Uganda as would be expected of a well-functioning national referral system. On a programming note, these data point to the importance of devising capacity building interventions for facilities persistently referring to KNRH such as tailoring remedial training of health workers in skills gaps revealed by the nature of incoming referrals (such as the need for refresher EMOnC and partograph trainings). In addition, there is need to develop a robust feedback mechanism to lower referring facilities in maternal care service provision such as in the proper management of cesarean sections which is the leading source of incoming referrals and in general, emergency obstetric management. Mentorship and support supervision of lower referring facilities in the provision of maternal care services in Uganda such as help in diagnosing complications in pregnancy is recommended.

Our study suggests a need for strengthening documentation of routine maternal and newborn care at the primary care and secondary level of service delivery in Uganda to support their appropriate management at the pinnacle of the national referral system which needs these referral data. This may call for a review of the health information systems (HMIS) in as far as it is supportive of a high quality of maternal and newborn services along the maternal care cascade.

In this study we found that there were a number of adolescent and child mothers among the incoming referrals to KNRH and that obstructed labour was the leading indication of referral for this group of mothers. Previous studies have highlighted teenage pregnancy as a risk factor for obstructed labor and related complications of early pregnancy [25]-[28]. With over 224 child mothers (<14 years) referred to KNRH over a period of 12 months, our findings suggest that early pregnancies could be common in Uganda and we call for policy and programming interventions aimed at reducing the burden of teenage pregnancies in Uganda and the broader region of sub-Saharan Africa [29]-[33].

## Data Availability

All data produced in the present study are available upon reasonable request to the authors

